# Sex-specific differences in nonlinear associations between glycaemia and brain health in UK Biobank

**DOI:** 10.64898/2026.03.12.26348052

**Authors:** Nasri Fatih, Sarah-Naomi James, Nishi Chaturvedi, Alun D Hughes, Victoria Garfield

## Abstract

**Aims:** Diabetes and hyperglycaemia are associated with poorer brain health, but most studies focus on diagnosed diabetes and rarely examine non-linear or sex-specific associations between glycaemia and neuroimaging biomarkers. We investigated whether glycaemia shows non-linear associations with brain structure and whether these differ by sex.

**Methods:** In 36,321 UK Biobank participants, sex-stratified fractional polynomial models assessed associations of random glucose and HbA1c with brain MRI volumes, adjusted for covariates. Restricted cubic splines and range-restricted linear models assessed robustness and estimated sex-specific slopes.

**Results:** Associations of glycaemia with whole brain and grey matter volumes were inverted J-shaped in both sexes, with lower volumes at higher glycaemia and less consistent evidence of lower volumes at the lower end. Predicted whole brain volume peaked at glucose values between 4.4 and 4.7 mmol/L and HbA1c values between 33 and 37 mmol/mol.

Restricted cubic splines confirmed non-linearity for glucose with whole brain volume (p_non-linearity=0.006) and grey matter (p_non-linearity<0.001), with evidence of sex interaction (p_interaction=0.02–0.03). For HbA1c, splines supported non-linearity for whole brain volume (p_non-linearity=0.006) and grey and white matter volumes (both p_non-linearity<0.001), with little evidence of sex interaction (p_interaction≥0.4). White matter hyperintensity volume showed a J-shaped association for HbA1c (p_non-linearity=0.002), with no evidence of sex interaction. Linear models in the descending range indicated steeper glucose-associated declines in whole brain and grey matter volumes in females than males.

**Conclusions:** Glycaemia–brain imaging associations were non-linear. Higher glycaemia was associated with less favourable brain structural profiles, with stronger associations for glucose in females. These findings indicate that associations between glycaemia and brain structure extend across the glycaemic spectrum rather than being limited to clinically defined diabetic states.

**What is already known about this subject?:**

- Diabetes is associated with adverse brain outcomes including reduced brain volume and greater small vessel disease burden and is an established risk factor for dementia.
- Most neuroimaging studies have focused on diabetes status rather than the full glycaemic continuum and have relied on sex-pooled analyses.
- Emerging evidence suggests glycaemia may relate to health outcomes nonlinearly and that these relationships may differ by sex.

**What is the key question?:**

- Is glycaemia associated with structural brain health across its full range, including below diabetes diagnostic thresholds, and do these associations show nonlinear or sex-specific patterns?

**What are the new findings?:**

- We found evidence of nonlinear associations between glycaemia and brain structure in both sexes, with inverted J-shaped associations for whole brain and grey matter volumes.
- Higher glycaemia was associated with less favourable brain structural profiles across the glycaemic continuum, including within the non-diabetic range.
- Sex differences were observed particularly for glucose, with steeper declines in brain volumes at higher glycaemic levels in females than males.

**How might this impact on clinical practice in the foreseeable future?:**

- These findings reinforce the importance of considering glycaemia as a continuous exposure rather than relying on diagnostic thresholds when evaluating metabolic contributions to brain ageing.

## Introduction

Diabetes mellitus is associated with poorer brain health and an increased risk of dementia. [1, 2] Some neuroimaging studies have also linked type 2 diabetes to reduced total brain volume, grey matter atrophy and increased white matter hyperintensity burden. [3] However, most neuroimaging studies have focused on diagnosed diabetes rather than examining glycaemia continuously and have typically relied on sex-pooled analyses, despite evidence that females with diabetes face a higher risk of adverse health outcomes than males, even though their overall risk of developing diabetes is lower.[4–6] Less is known about sex differences in the relationship between glycaemia spanning the normal and pathological range and brain health in population-based samples; our previous analyses in a British birth cohort found that the association between higher HbA1c levels, insulin resistance and glucose and lower whole brain volume (WBV) in older age was stronger in females than males. [7, 8]

Other studies in large cohorts have demonstrated nonlinear associations between glycaemia and a range of health outcomes. For cardiovascular disease, HbA1c has shown a nonlinear relationship with risk across the glycaemic spectrum.[9] Similarly, nonlinear bell-shaped and U-shaped associations have been reported between HbA1c and cognitive impairment in alcohol use disorder and bone mineral density in postmenopausal women with type 2 diabetes respectively.[9–11] Collectively these findings suggest that assuming linearity between glycaemia and health outcomes may be an oversimplification, and that the shape of association may vary by outcome and population — raising the question of whether similar nonlinear patterns exist for brain structure. The current analyses adopted an a priori sex-stratified and nonlinear approach to examine the relationship between HbA1c and random glucose with neuroimaging markers. HbA1c and random glucose capture different aspects of glycaemic physiology — chronic average exposure versus acute fluctuation — and may therefore relate differently to brain structural outcomes. [12] UK Biobank provides a unique opportunity to investigate these associations across the full glycaemic spectrum, including levels below diagnostic thresholds for diabetes. Understanding these patterns may help clarify whether brain-relevant glycaemic variation operates across the full population range rather than only at clinically defined extremes. We hypothesised that glycaemic markers would show non-linear associations with brain health, and that these associations may differ by sex.

## Methods

### Sample

We used data from participants in UK Biobank, a cohort of ∼500,000 individuals (94% self-reported European ancestry) aged 40–69 at baseline, recruited between 2006 and 2010. Baseline assessments included questionnaires, interviews, biological samples, and linkage to routine health data. Since 2014, a subsample of 52,951 has undergone brain imaging by MRI at dedicated centres. Inclusion criteria for the current analysis were: imaging data available at the time of extraction, at least one measure of glycaemia, no evidence of dementia at baseline, and self-reported White European ethnicity. Reporting conformed to STROBE guidelines [13]

### Variables

#### Exposures

HbA_1c_ was measured from whole blood using Bio-Rad Variant II Turbo analysers (HPLC method), with calibration ensured via multi-instrument comparison [14]. Random glucose was measured from serum using hexokinase analysis on a Beckman Coulter AU5800.

Confounders: these were chosen based on prior evidence and informed by directed acyclic graphs. We adjusted for age, educational attainment (years of full-time education, based on International Standard Classification of Education [ISCED]), socioeconomic deprivation (Townsend index quintiles), body mass index (weight [kg]/height[m]^2^), and smoking status (never, former, current). Education and smoking status were derived from baseline questionnaires; BMI was measured at the assessment centre; and Townsend deprivation index was derived from census data linked to participants’ postcodes.

#### Outcomes

Brain MRI was conducted using standard protocols (discussed in Supplementary Material). Imaging outcomes included hippocampal volume (HV) and white matter hyperintensity volume (WMHV), both adjusted for total intracranial volume (TIV), and whole brain volume (WBV), grey matter (GM), and white matter (WM), normalised for head size.

#### Sex

Sex was analysed as recorded in UK Biobank. It was derived from central registry records at recruitment and could be updated by participants; thus, the variable reflects registry-recorded sex with possible participant updates.

No matching was performed; confounding was handled via multivariable adjustment (Models 1–2) and sex-stratified analyses.

### Statistical analysis

Analyses were performed using Stata 18.0 and derived variable list are included in the Supplementary Material. Continuous variables were summarised as mean (SD) or median [IQR], as appropriate. White matter hyperintensity volume (WMHV) was log-transformed due to positive skew. Fractional polynomial (FP) regression was used to flexibly model potential non-linear associations between glycaemic markers (HbA1c and random glucose) and brain imaging measures, allowing up to two FP powers selected from the set {−2, −0.5, 0, 0.5, 1, 2, 3}[15]. Model fit was evaluated using deviance statistics, and the most parsimonious FP model was selected as the lowest-degree model that was not rejected when compared with the model with the minimum deviance.

A priori, sex-stratified analyses were conducted using two adjustment levels: Model 1 included age (and total intracranial volume [TIV] where appropriate), and Model 2 additionally adjusted for education, deprivation, smoking status, and body mass index (BMI). Sensitivity analyses excluded (i) participants using hypoglycaemic medication and (ii) individuals with diabetes, to assess the robustness of associations to treatment effects and diagnosed disease status.

To assess robustness, FP results were compared with restricted cubic spline (RCS) models using five knots placed at the 5th, 27.5th, 50th, 72.5th, and 95th percentiles of the exposure distribution.

When non-linearity was observed, we fitted additional linear models within the ascending and descending portions of the curve to estimate sex-specific slopes, using ranges defined from the fitted non-linear plots. Sex differences in spline associations were assessed in pooled models including sex-by-spline interaction terms.

To further characterise sex differences in curve shape, we examined spline-derived peak derivative metrics. Specifically, *we identified key turning points from the spline function, including the peak (x,* ŷ) and the onset of decline (x□□), and quantified the point of maximum positive gradient (x(dmax), ŷ(dmax)), which captures where outcomes increase most rapidly across the ascending portion of the curve. From this analysis, x* and ŷ* denote the glycaemia value and corresponding predicted outcome at the peak of the curve; x(dmax) and ŷ(dmax) represent the glycaemia value and predicted outcome at the point of maximum positive gradient; and x□□ indicates the onset of decline, defined as the point where the slope transitions from positive to negative. These coordinates were extracted separately for males and females to facilitate comparison of sex-specific exposure–response profiles.

## Results

A total of 36,321 participants were included (mean age 55y; 47% males – see flow diagram in Figure 1). Sample characteristics stratified by sex are presented in Table 1. Males were slightly older than females, had higher HbA_1c_ and random glucose levels. They were also more likely to be current or ex-smokers, to have a diabetes diagnosis and had higher whole brain volume.

**Figure 1:**
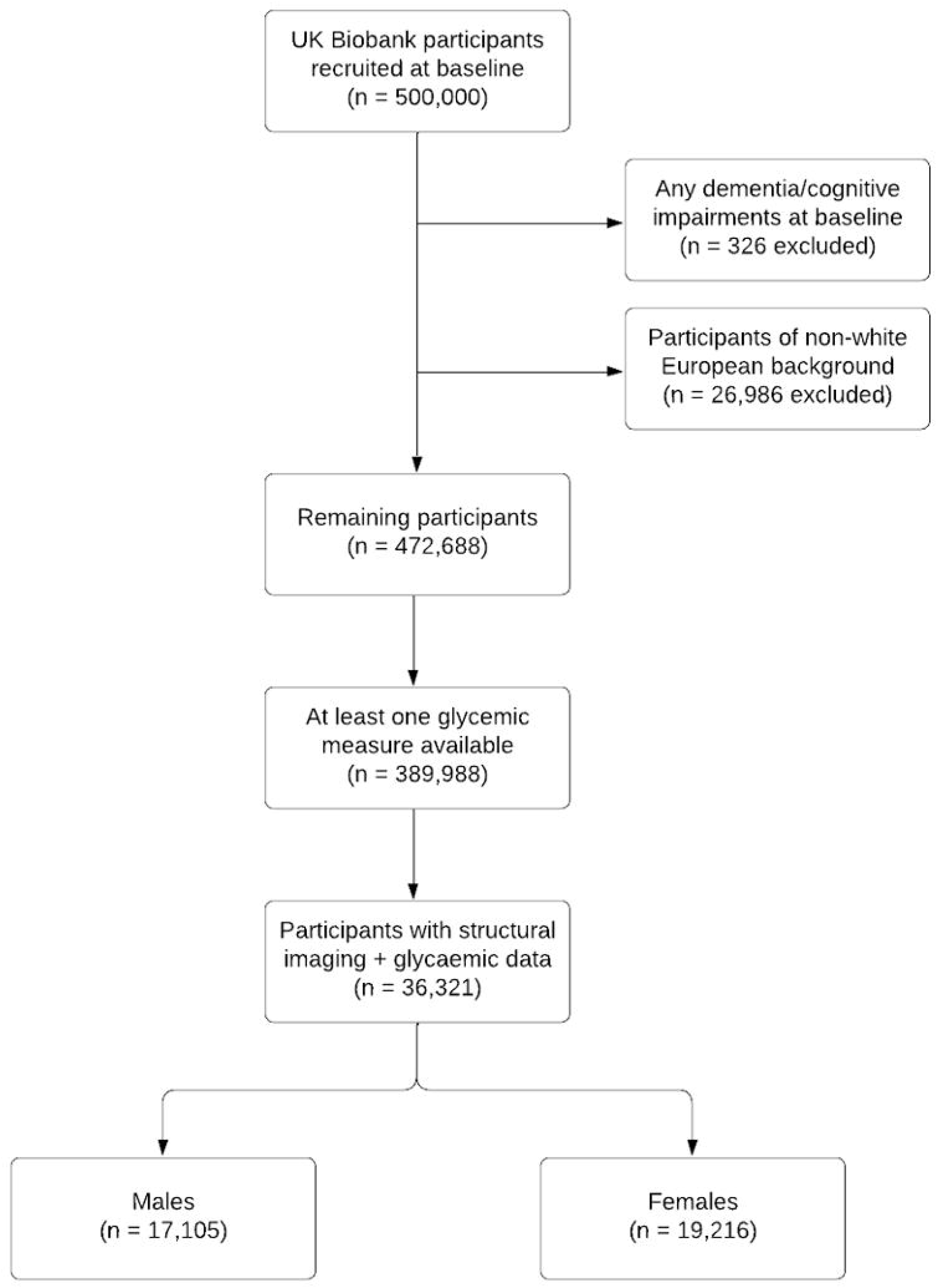
Flowchart of participant inclusion.

**Table 1:**
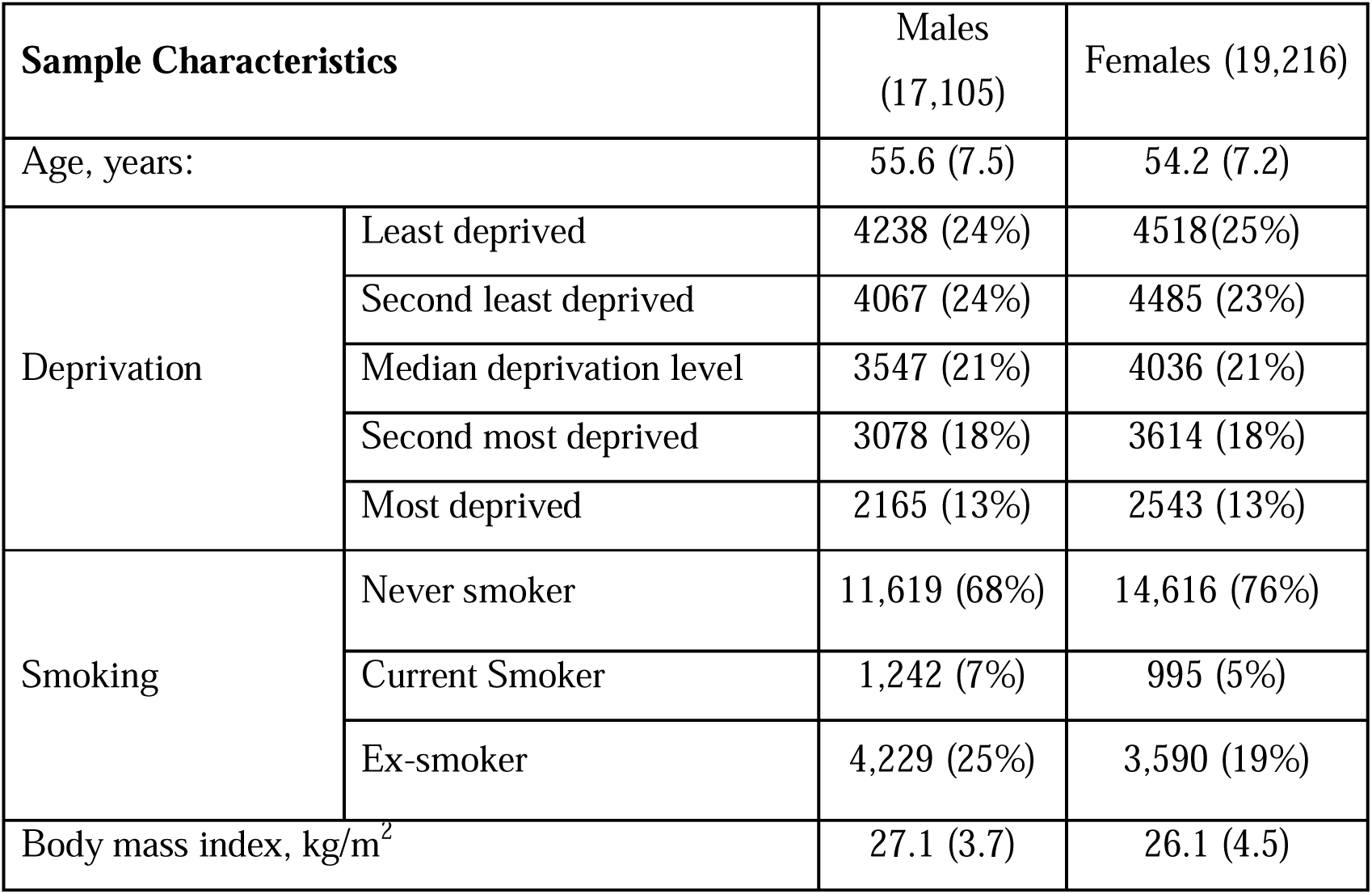
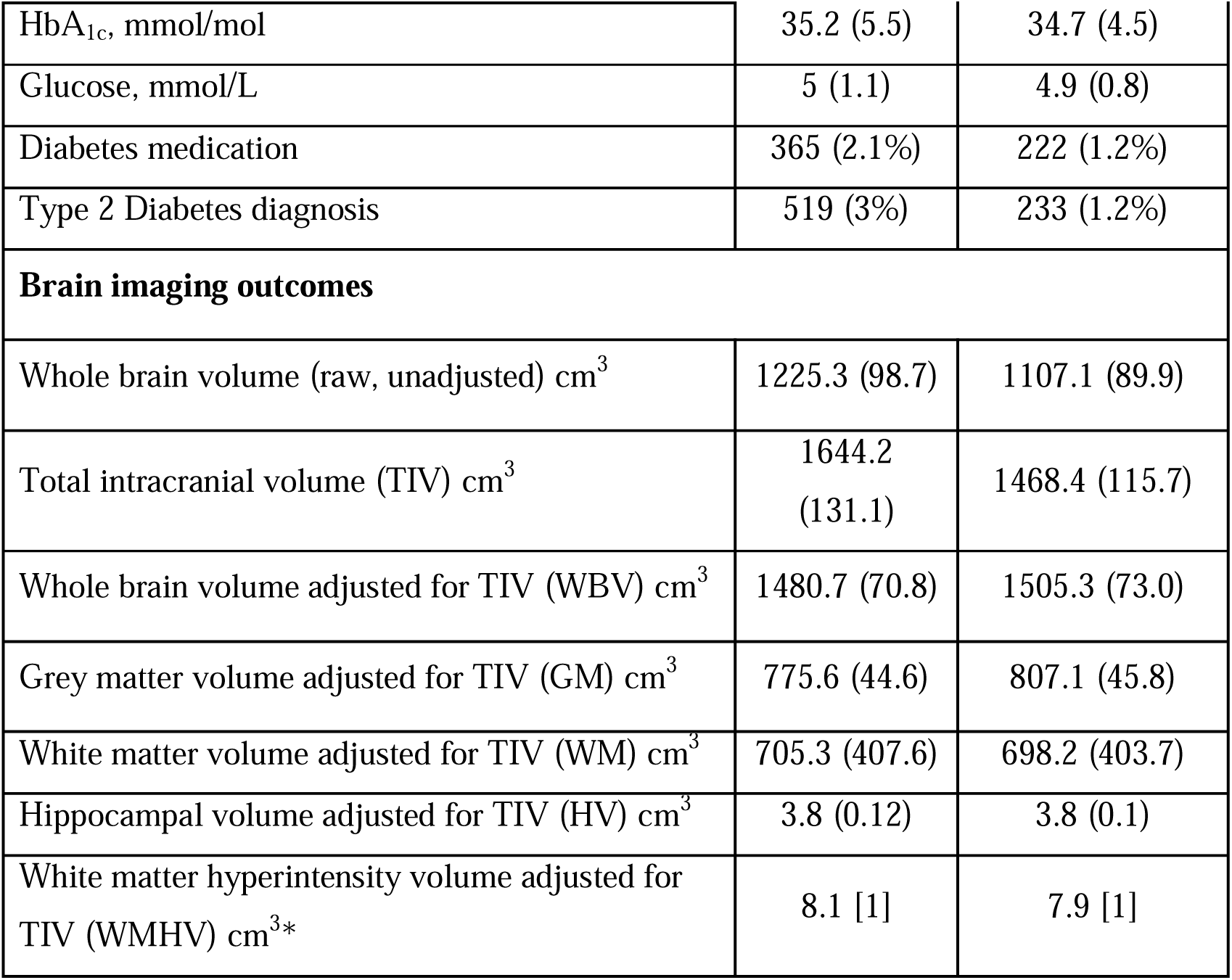
Sample characteristics of UK Biobank participants included in the analysis, overall and by sex. . Data are presented separately for men and women. Continuous variables are shown as mean (SD) and categorical variables as n (%), unless otherwise specified. Brain volumes are presented as unadjusted values and as volumes adjusted for total intracranial volume (TIV). White matter hyperintensity volume was additionally adjusted for TIV and log-transformed where appropriate for analysis. Deprivation was categorised into quintiles from least to most deprived. Diabetes medication refers to self-reported use or recorded prescription of glucose-lowering medication. WBV=whole brain volume. GM=grey matter volume. WM=white matter volume. HV=hippocampal volume. WMHV=white matter hyperintensity volume.

| Sample Characteristics |  | Males<br>(17,105) | Females (19,216) |
| --- | --- | --- | --- |
| Age, years: |  | 55.6 (7.5) | 54.2 (7.2) |
| Deprivation | Least deprived | 4238 (24%) | 4518(25%) |
|  | Second least deprived | 4067 (24%) | 4485 (23%) |
|  | Median deprivation level | 3547 (21%) | 4036 (21%) |
|  | Second most deprived | 3078 (18%) | 3614 (18%) |
|  | Most deprived | 2165 (13%) | 2543 (13%) |
| Smoking | Never smoker | 11,619 (68%) | 14,616 (76%) |
|  | Current Smoker | 1,242 (7%) | 995 (5%) |
|  | Ex-smoker | 4,229 (25%) | 3,590 (19%) |
| Body mass index, kg/m<sup>2</sup> |  | 27.1 (3.7) | 26.1 (4.5) |

|  |  |  |
| --- | --- | --- |
| HbA<sub>1c</sub>, mmol/mol | 35.2 (5.5) | 34.7 (4.5) |
| Glucose, mmol/L | 5 (1.1) | 4.9 (0.8) |
| Diabetes medication | 365 (2.1%) | 222 (1.2%) |
| Type 2 Diabetes diagnosis | 519 (3%) | 233 (1.2%) |
| **Brain imaging outcomes** |  |  |
| Whole brain volume (raw, unadjusted) cm<sup>3</sup> | 1225.3 (98.7) | 1107.1 (89.9) |
| Total intracranial volume (TIV) cm<sup>3</sup> | 1644.2<br>(131.1) | 1468.4 (115.7) |
| Whole brain volume adjusted for TIV (WBV) cm<sup>3</sup> | 1480.7 (70.8) | 1505.3 (73.0) |
| Grey matter volume adjusted for TIV (GM) cm<sup>3</sup> | 775.6 (44.6) | 807.1 (45.8) |
| White matter volume adjusted for TIV (WM) cm<sup>3</sup> | 705.3 (407.6) | 698.2 (403.7) |
| Hippocampal volume adjusted for TIV (HV) cm<sup>3</sup> | 3.8 (0.12) | 3.8 (0.1) |
| White matter hyperintensity volume adjusted for TIV (WMHV) cm<sup>3</sup>* | 8.1 [1] | 7.9 [1] |

### Fractional polynomial models

Relationships between glycaemic markers (glucose and HbA1c) and brain volume measures (WBV, GM, and WM) were non-linear (inverted J-shaped) in both sexes (Figures 2–3). A two-term fractional polynomial model provided the best fit for whole brain volume in both males and females, although the selected powers differed by sex (Supplementary Table 1).

**Figure 2:**
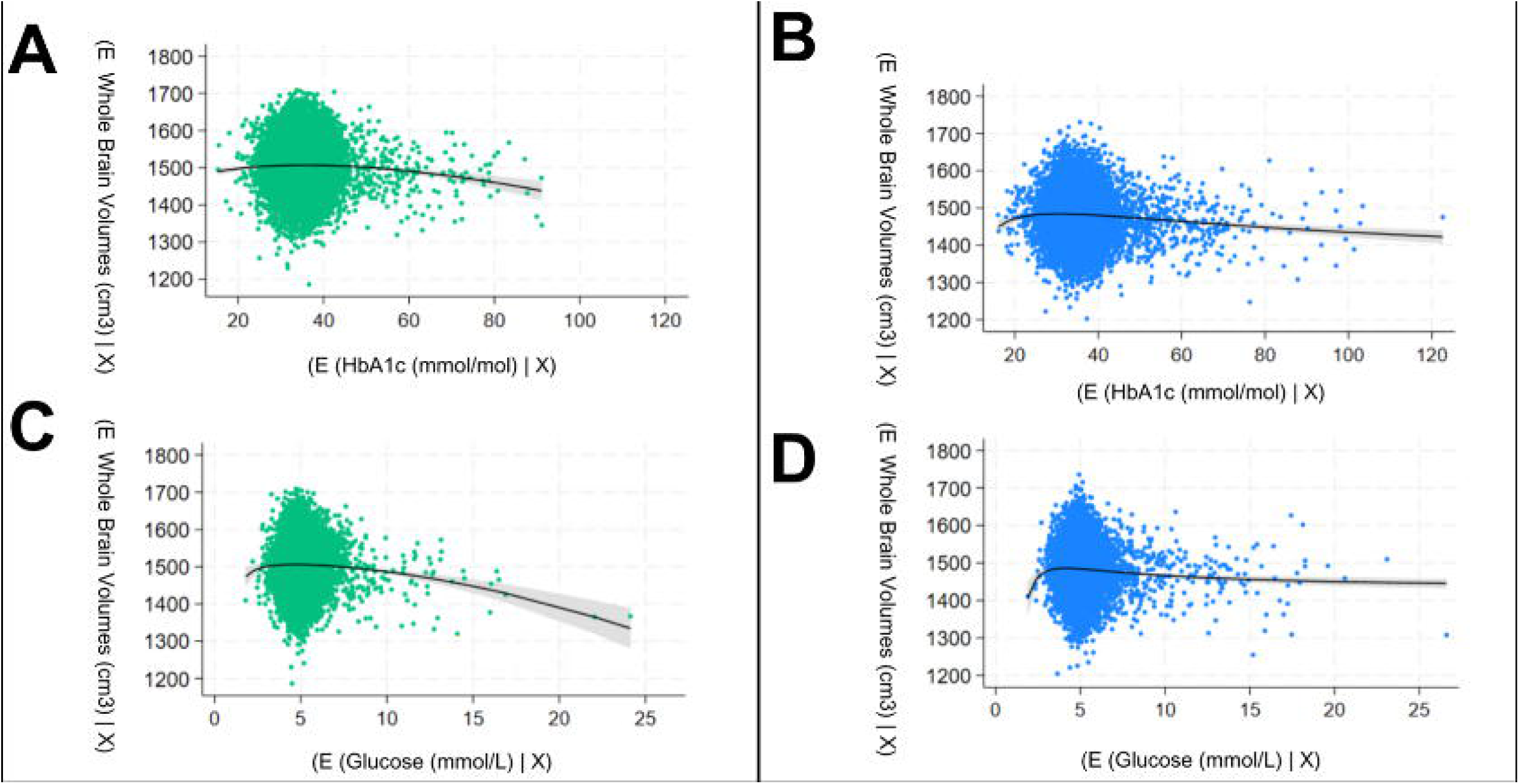
Multivariable fractional polynomial models with 95% confidence limits of the relationship between HbA_1c_ (A and B) and glucose (C and D). and whole brain volumes. Females are represented in green (A and C) and males in blue (B and D). E(Y) on the y-axis represents the predicted values of the response variable, while E(X|Xn) on the x-axis represents the conditional expected values of the predictor variable of interest.

**Figure 3:**
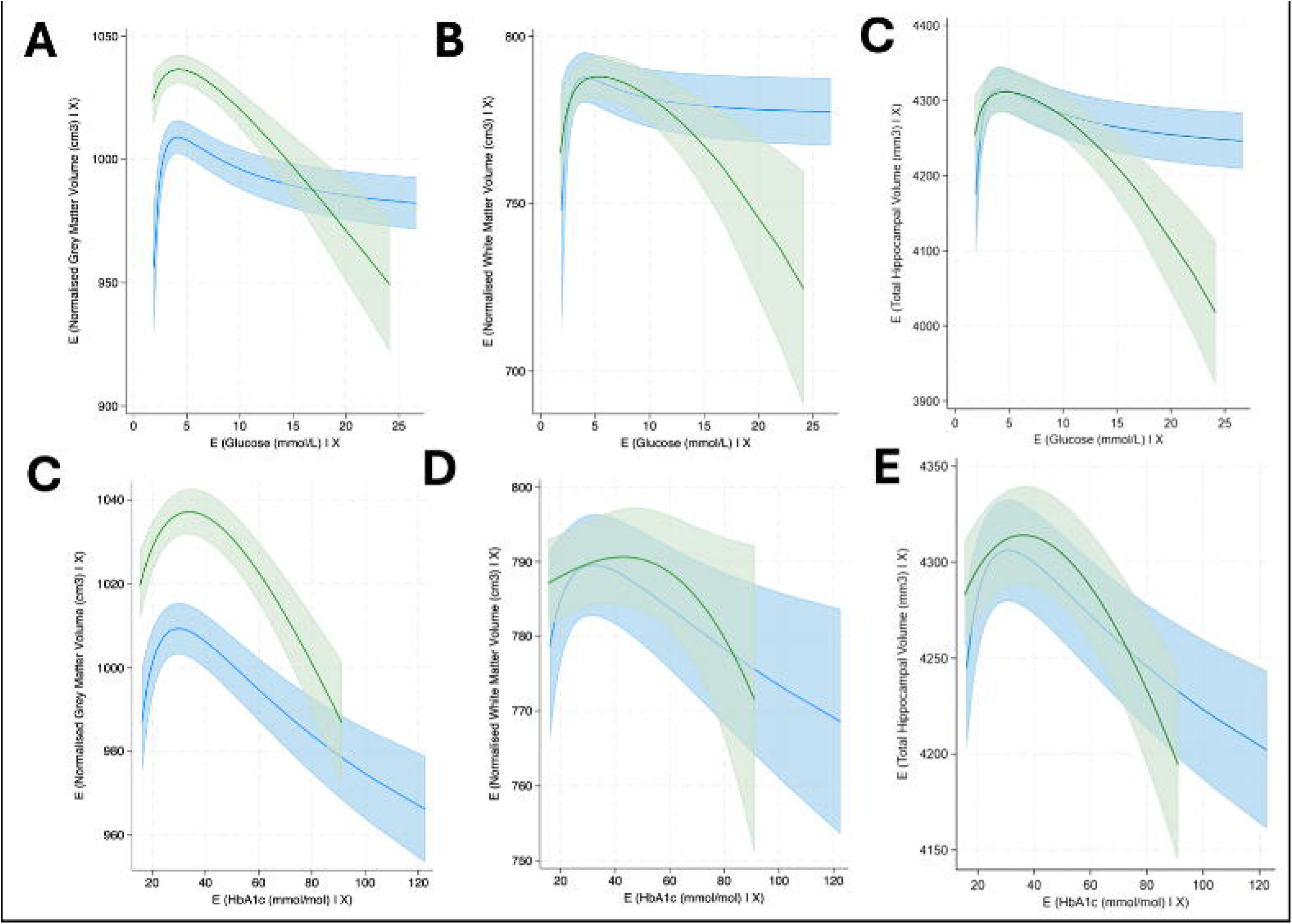
Partial regression plots of the relationship between glucose and (A) grey matter, (B) white matter volumes, (C) hippocampal volume; or HbA1c and (D) grey matter, (E) white matter, (F) hippocampal volume. Fractional polynomial curves are shown for the fully-confounder adjusted models and individual data points have been omitted for clarity. Relationships for females are shown in green and males in blue. Other abbreviations as for Figure 2.

Visually, the decline in brain volumes at higher glycaemia (above 4.5 mmol/L glucose or ∼40 mmol/mol HbA1c) appeared steeper in females than males, although for HbA1c the GM and WM curves largely overlapped between sexes, with wider uncertainty at the extremes indicating less precise sex-specific estimates. WMHV also showed a J-shaped pattern, most clearly for HbA1c, with no clear evidence that the curve shape differed by sex.

### Restricted cubic splines

Findings were similar using restricted cubic splines, which confirmed non-linear associations for glucose with WBV (p_non-linearity = 0.006) and GM (p_non-linearity < 0.001), with evidence of sex interaction for both outcomes (p_interaction = 0.02 and 0.03, respectively). For HbA1c, spline models also supported non-linearity for WBV (p_non-linearity = 0.006), GM and WM (both p_non-linearity < 0.001), with little evidence of sex interaction (p_interaction ≥ 0.4). Consistent with the FP models, associations increased across the lower range before declining at higher levels; results were robust to alternative knot placements. For HV, spline models showed no evidence of non-linearity for either glucose or HbA1c; however, there was evidence of sex interaction for HbA1c–HV (p_interaction = 0.0001) but not for glucose–HV (p_interaction = 0.79). For WMHV, spline models supported non-linearity for HbA1c (p_non-linearity = 0.002) but not glucose, with no evidence of sex interaction for either marker. The plots are represented in the appendix (Figure 4).

**Figure 4:**
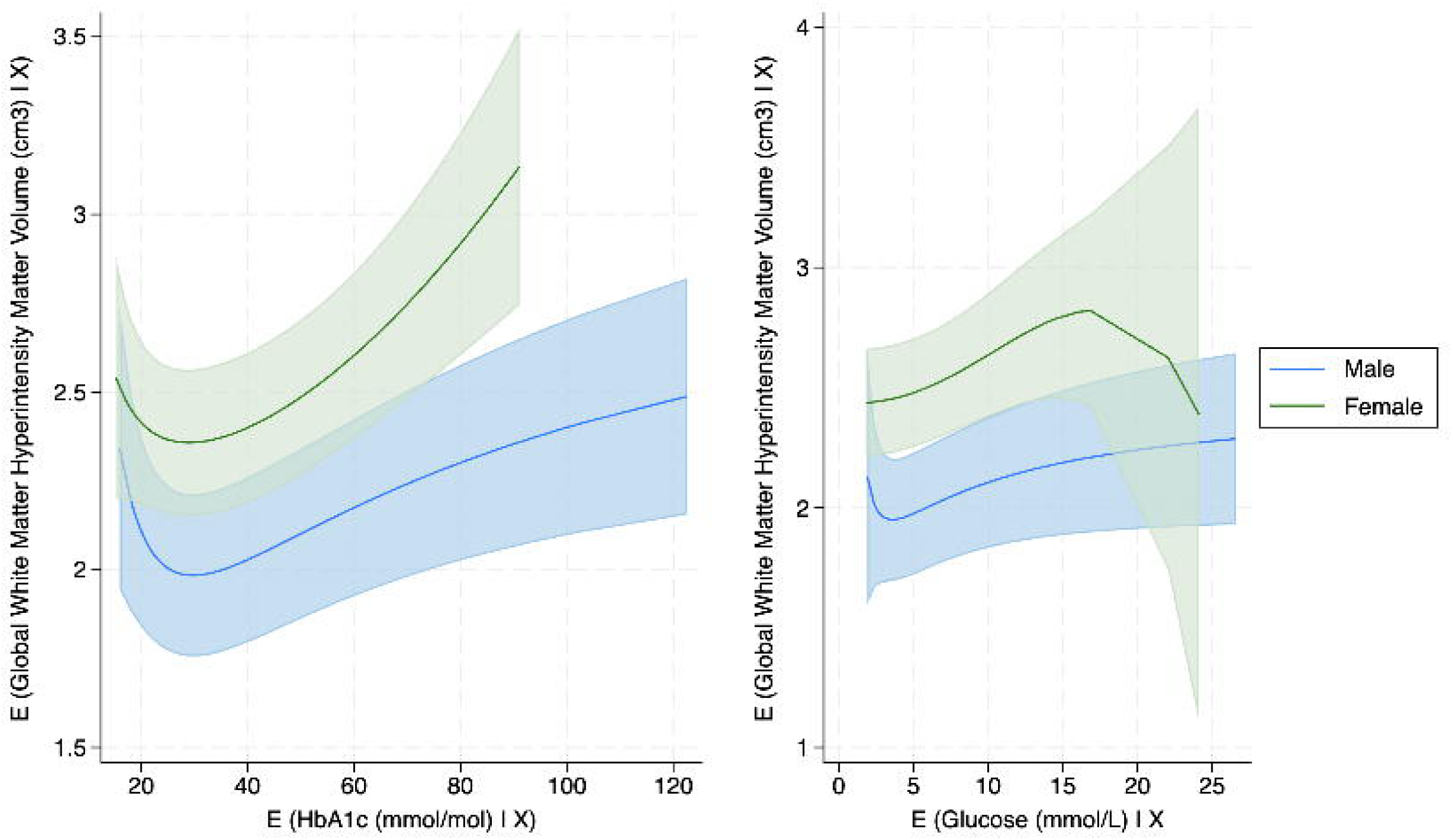
Partial regression plots for the fully confounder-adjusted relationships between HbA_1c_ and glucose on white matter hyperintensity volumes. Females in green, males in blue. Other abbreviations as for Figure 3.

### Peak derivatives

For glucose, WBV and GM showed non-linear, threshold-dependent associations. Predicted WBV peaked at 4.7 mmol/L in males (x* = 4.7; 95% CI 3.6–5.8; ŷ = 1489.4 cm³) and 4.4 mmol/L in females (x* = 4.4; 95% CI 3.8–5.0; ŷ = 1505.9 cm³), with volumes declining beyond ∼5 mmol/L. GM showed a similar pattern, peaking at 4.78 mmol/L in males (95% CI 3.69–5.86; ŷ = 782.6 cm³) and 4.40 mmol/L in females (95% CI 3.00–5.81; ŷ = 807.3 cm³), with an earlier onset and steeper post-peak decline in females. WMHV also showed a J-shaped pattern, with a sharp increase beyond ∼5 mmol/L; the maximum-slope point occurred at ∼5.1 mmol/L in both sexes (males: xdmax = 5.13; females: xdmax = 5.15), and predicted WMHV peaked at 26.6 mmol/L (males: ŷ = 8.49 cm³; females: ŷ = 8.43 cm³).

For HbA1c, WBV peaked at 37.4 mmol/mol in females (95% CI 20.0–54.8; ŷ = 1504.6 cm³) and 33.2 mmol/mol in males (95% CI 27.4–39.0; ŷ = 1489.7 cm³), with volumes declining above these thresholds. GM peaked at 31.8 mmol/mol in females (95% CI 17.5–46.2; ŷ = 806.4 cm³) and 34.2 mmol/mol in males (95% CI 22.9–45.4; ŷ = 782.0 cm³), with decline beginning around ∼32–34 mmol/mol. WMHV also increased non-linearly with HbA1c, peaking at 122.6 mmol/mol in both sexes (males: ŷ = 8.70; females: ŷ = 8.68); the maximum-slope point occurred at 36.7 mmol/mol in males and 108.4 mmol/mol in females.

### Linear models

In linear models restricted to participants with high glucose (>10 mmol/L)—a range corresponding to the descending portion of the fitted non-linear association where relationships appeared approximately linear—there was evidence of sex differences in the strength of association between glucose and WBV and GM, with steeper declines in females than males (WBV: β_females = −7.22 cm³ per mmol/L [95% CI −14.1, −0.34] vs β_males = −1.7 cm³ [−6.4, 2.9]; p_interaction = 0.05; GM: β_females = −4.1 cm³ [−8.2, −0.1] vs β_males = 0.02 cm³ [−2.5, 2.6]; p_interaction = 0.04). There was no evidence of sex interaction for glucose–WMHV (p_interaction = 0.9) or glucose–HV (p_interaction = 0.94). In linear models restricted to HbA1c >60 mmol/mol (descending range), there was no evidence of sex interactions across outcomes (all p_interaction ≥ 0.1).

Across the lower (“uptick”) range, linear estimates for glucose were generally small and imprecise, with no convincing evidence of sex differences for HV (p_interaction = 0.20) or WMHV (p_interaction = 0.28). For HbA1c in the uptick range (20–42 mmol/mol), there was evidence of sex interaction for both HV (p_interaction < 0.001) and WMHV (p_interaction = 0.018). Across both glucose and HbA1c, linear models fitted within the ascending and descending portions provided little evidence of sex differences for WM (all p_interaction ≥ 0.5), and the estimated slopes were generally small and imprecise.

These are presented in Supplementary Table 2:

#### Sensitivity analyses

Sensitivity analyses using fractional polynomial models excluding individuals who were either on diabetes medication or had diabetes were not materially different (see Supplementary Figure S1 and Figure S2).

## Discussion

### Statement of principal findings

We observed non-linear inverted J-shaped relationships between both HbA1c and random glucose and brain volume measures in a large population-based sample from UK Biobank; higher glycaemia — and possibly lower levels at the extremes — were associated with lower volumes, consistent with less favourable brain profiles. These non-monotonic associations were present in both sexes, with females showing a somewhat steeper decline at higher glycaemic levels, particularly for glucose. Excluding individuals with diagnosed diabetes or on glucose-lowering medication did not materially change the overall pattern. Together, these findings emphasise the importance of analysing glycaemia as a continuous exposure and highlight the complexity—and possible sex dependence—of its relationship with neuroimaging markers of brain health. The differing temporal characteristics of HbA1c and random glucose may also capture distinct physiological processes, which could partly explain differences in the strength and consistency of observed associations across outcomes.

Across modelling approaches (fractional polynomials, restricted cubic splines, and range-restricted linear models), results were largely concordant: non-linearity was most consistently supported for glucose and HbA1c with WBV and GM (and for HbA1c with WM), while sex differences were selective rather than universal across outcomes—strongest for glucose with WBV and GM, and less evident for HbA1c across most outcomes. WMHV showed a J-shaped pattern most clearly for HbA1c, whereas evidence for a comparable glucose–WMHV non-linearity was weaker, and there was little support for sex differences in WMHV. Although peak-derivative analyses helped characterise putative thresholds around which associations changed direction, estimates at the extremes were imprecise, so apparent turning points and sex differences should be interpreted cautiously as descriptive rather than definitive.

### Strengths and weaknesses in relation to other studies, discussing important differences in results

The finding that glycaemia, namely, HbA1c shows non-linear associations with several health outcomes has been reported previously [9],[10],[11]; however, evidence relating HbA1c to brain health is more limited. A Japanese clinical study reported a non-linear (U-shaped) association between HbA1c and the presence of white matter lesions in females.[16] In contrast, a previous UK Biobank study categorised HbA1c into five clinical groups and reported that low-normal HbA1c was associated with higher hippocampal volume in pooled analyses of males and females. [17] We did not observe a consistent low-normal benefit in our analyses when glycaemia was modelled as a continuous exposure.

This difference may partly reflect methodological choices: pooling sexes can mask effect modification, and categorising a continuous exposure may shift or obscure underlying non-linear relationships by imposing artificial thresholds, particularly around turning points in the exposure–response curve. Our observation that elevated glycaemia above the normoglycaemic range is associated with adverse brain health is consistent with previous population-based studies and studies in people with T2D [7, 18, 19]. More broadly, emerging work using flexible modelling approaches has also reported non-linear associations between glycaemic metrics and structural or cognitive brain outcomes, suggesting that the shape of the relationship may depend on how glycaemia is parameterised.

Multiple potential mechanisms have been postulated to account for the association between elevated glycaemia and adverse brain structural outcomes. Chronic hyperglycaemia may promote oxidative stress and neuroinflammation, contributing to neuronal damage and grey matter loss.[20] Cerebrovascular effects — including endothelial dysfunction, impaired cerebral autoregulation and small vessel disease — may underlie associations with white matter hyperintensity burden in particular [21] Additionally, hyperglycaemia has been linked to impaired amyloid clearance and altered neuronal insulin signalling, both of which may contribute to accelerated brain ageing. [22] The relative contribution of these pathways likely varies across the glycaemic spectrum, which may partly explain why associations with brain structure appear non-linear rather than uniformly dose dependent.

Possible associations between lower glycaemic levels and brain health have received comparatively little attention, except in the context of severe hypoglycaemia due to insulin treatment or endocrine abnormalities [23]. Previous studies have shown that hypoglycaemic events in those with T2D can be detrimental to the brain, increasing the risk of dementia and cognitive decline[23, 24], although these studies often include states of considerably lower glucose than observed in this study. Hypoglycaemia can be associated with a range of negative neurophysiological consequences such as oxidative stress, microgliosis, impaired synaptic plasticity and neuronal death [24–26]. We previously reported that higher HbA_1c_ was more strongly associated with reduced whole brain and grey matter volumes in females than males in a British birth cohort [7]. In the present study, sex differences were most evident for glucose at higher glycaemic levels, while for HbA1c the fitted GM and WM curves were largely overlapping between sexes with limited evidence of interaction. Because the apparent strength of a linear association can be influenced by the exposure distribution within each sex, underlying non-linear relationships may also create the impression of sex differences even when overall curve shapes are similar. These findings highlight the importance of accounting for non-linearity when evaluating sex differences in glycaemia–brain imaging associations.

#### Strengths and weaknesses of the study

Our study has several strengths. First, UK Biobank is one of the largest population-based cohorts with both glycaemic markers—capturing short-term and longer-term glycaemia—and detailed neuroimaging measures. Using both random glucose and HbA1c, each with distinct strengths and limitations [27], increases confidence in the robustness of the observed non-linear associations. The large sample size enabled evaluation of glycaemia across its full population distribution, including potential non-monotonic relationships with brain measures, and supported subgroup analyses such as sex-stratified models. Finally, deep baseline phenotyping allowed adjustment for a range of sociodemographic, lifestyle, and metabolic covariates, reducing the risk of confounding. A further strength is the use of complementary modelling approaches to characterise non-linearity, including fractional polynomials as the primary framework with restricted cubic splines as a robustness check. We also quantified key curve features using spline-derived turning points and estimated sex-specific slopes within approximately linear regions of the exposure–response relationship.

Nevertheless, our study has limitations. Its cross-sectional design limits causal inference, and UK Biobank is not fully representative of the general population, with potential for healthy volunteer selection and survival biases. These factors may influence the magnitude of associations and, to a lesser extent, the apparent shape of non-linear relationships. Additionally, lower glycaemic levels may reflect underlying physiological or health states rather than a relationship with brain structure, raising the possibility of reverse causation at the lower end of the exposure distribution. The restriction to a largely White European cohort further limits generalisability to more diverse populations in whom glycaemic physiology and vascular risk profiles may differ.

### Meaning of the study: possible explanations and implications for clinicians and policymakers

These findings suggest that glycaemia–brain health relationships are unlikely to be adequately captured by conventional threshold-based approaches. Associations were evident across the full glycaemic spectrum rather than confined to clinically defined diabetic states, implying that risk may operate along a continuum, rather than emerging only beyond diagnostic thresholds.

The observed non-linear patterns raise the possibility that higher glycaemic levels — and possibly lower levels at the extremes — may relate to structural brain differences, although interpretation at the extremes remains cautious. From a clinical and public health perspective, these results reinforce the importance of considering glycaemic exposure dimensionally rather than categorically when evaluating potential neurological risk.

Longitudinal studies are needed to establish whether these associations reflect causal pathways, and should incorporate sex-stratified analyses and flexible modelling approaches to adequately capture the complexity of glycaemia–brain relationships across the life course.

## Conclusions

The associations between glycaemic markers and brain imaging outcomes in UK Biobank were non-linear in both males and females. For both HbA1c and glucose, whole-brain and grey-matter volumes were lower at higher glycaemia, with less consistent evidence of lower volumes at the lower end of the glycaemic distribution. WMHV showed evidence of a J-shaped association most clearly for HbA1c, with weaker evidence for comparable non-linearity for glucose. Together, these findings support a non-linear relationship between glycaemia and brain health and reinforce the value of modelling glycaemia as a continuous exposure rather than relying on categorical thresholds.

## Authors’ relationships and activities

NC receives funds from AstraZeneca to serve on data safety and monitoring committees for clinical trials. All other authors declare that there are no relationships or activities that might bias, or be perceived to bias, their work.

## Contribution statement

NF, AH, and VG conceived the study. AH provided statistical support. All authors interpreted the data. NF drafted the initial manuscript. All authors contributed to critical revision and editing of the manuscript. NF and AH accessed and verified the underlying data. The underlying data were verified by multiple co-authors (Victoria Garfield, Sarah-Naomi James, and Alun Hughes), and all authors agree to be accountable for all aspects of the work.

## Data availability

This research was conducted using the UK Biobank Resource under Application Number 7661. UK Biobank data are available to bona fide researchers through application via the UK Biobank Access Management System (https://www.ukbiobank.ac.uk). Individual-level data cannot be shared by the authors but can be accessed directly through UK Biobank upon approval. No new primary participant data were collected for this analysis.

## Code availability

The statistical code used for the analyses will be made available on GitHub upon publication.

## Ethics approval

UK Biobank has approval from the North West Multi-centre Research Ethics Committee (REC reference 11/NW/0382), and all participants provided written informed consent. This research was conducted under UK Biobank Application Number 7661.

## Consent to participate

Participants provided informed consent at the time of enrolment into UK Biobank.

## Consent for publication

Not applicable.

