## Supplementary File 1 for "Sex-specific differences in nonlinear associations between glycaemia and brain health in UK Biobank"

**Supplementary methods:**

**Details of the UK biobank neuroimaging protocol**

Participants were scanned at four imaging centres located in Central, North, South-East, and South-West England. A central team oversaw training and quality assurance across all sites, and all staff received extensive instruction from an MR physicist.

To ensure cross-site harmonisation, identical scanner models, software versions, calibration and tuning procedures, coil types, and imaging protocols were used throughout. Radiographers at each centre completed a standardised training programme, and standard operating procedures were implemented uniformly. Regular phantom scans, servicing, and performance checks were conducted under the supervision of a UK Biobank physicist.

Finally, both qualitative and quantitative image assessments were carried out by independent imaging experts to verify data quality and confirm suitability for research analyses.

**Supplementary table 1:** Fractional polynomial (FP) model comparison for HbA1c and glucose, stratified by sex. Models were fitted sequentially (omitted → linear → m = 1 → m = 2) and compared using deviance differences and likelihood ratio tests. The selected functional form (final powers shown in the “m = 2” row) reflects the best-fitting transformation based on deviance reduction and statistical significance.

|  | **HbA1c** | | | | | |
| --- | --- | --- | --- | --- | --- | --- |
|  | **Model** | **Test df** | **Deviance** | **Deviance Diff.** | **p** | **Powers** |
| **Males** | Omitted | 4 | 181408.4 | 52.062 | p <0.001 |  |
|  | Linear | 3 | 181370.7 | 14.353 | p <0.001 | 1 |
|  | m = 1 | 2 | 181367.3 | 11.028 | p <0.001 | 2 |
|  | m = 2 | 0 | 181356.3 | 0 | N/A | -2.5 |
| **Females** | Omitted | 4 | 203295.12 | 60.054 | p <0.001 |  |
|  | Linear | 3 | 203287.83 | 60.044 | p <0.001 | 1 |
|  | m = 1 | 2 | 203275.82 | 60.025 | 0.005 | 2 |
|  | m = 2 | 0 | 203265.34 | 60.009 | N/A | -2.5 |
|  | **Glucose** | | | | | |
|  | **Model** | **Test df** | **Deviance** | **Deviance Diff.** | **p** | **Powers** |
| **Males** | Omitted | 4 | 165592.91 | 41.672 | p <0.001 |  |
|  | Linear | 3 | 165570.07 | 18.833 | p <0.001 | 1 |
|  | m = 1 | 2 | 1655557.4 | 6.197 | p <0.04 | 3 |
|  | m = 2 | 0 | 165551.24 | 0 | N/A | -2 2 |
| **Females** | Omitted | 4 | 145546.55 | 60.907 | p <0.001 |  |
|  | Linear | 3 | 145494.44 | 60.788 | p <0.001 | 1 |
|  | m = 1 | 2 | 145494.44 | 60.788 | 0.005 | 1 |
|  | m = 2 | 0 | 145484.16 | 60.767 | N/A | -3 |

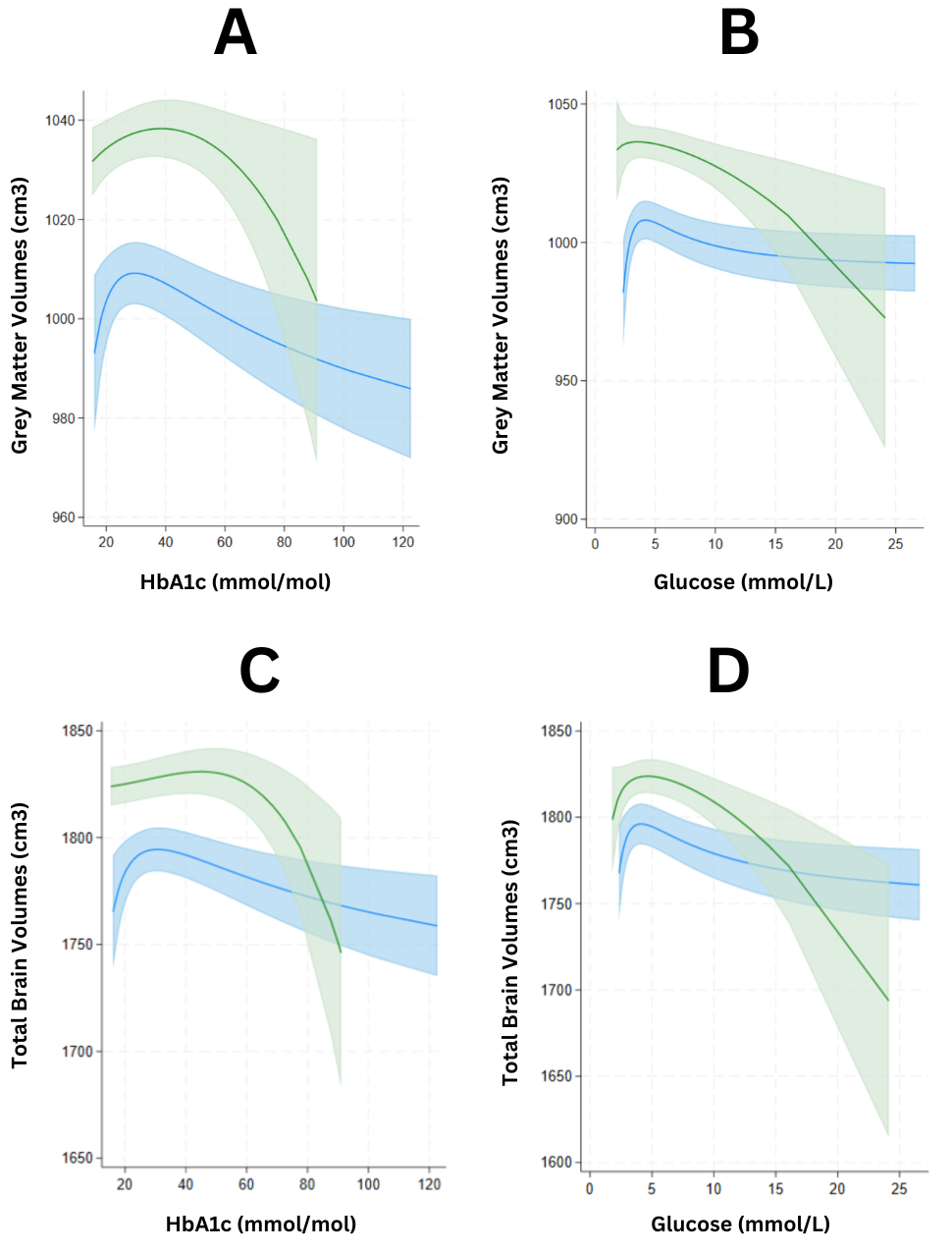

**Supplementary figure 1:** The partial regression plot of the fractional polynomial models that provided the best fit for the relationship between HbA_1c_/glucose and grey matter volume(A and B) and whole brain volume (C and D). The models presented are the fully confounder-adjusted models excluding those on diabetes medication.

These relationships for females are presented in green and for the males in blue. Confidence limits are represented by the shade surrounding the line. As per the plot above, the predicted values of the response variable and the conditional expected values of the predictor variable of interest are displayed.

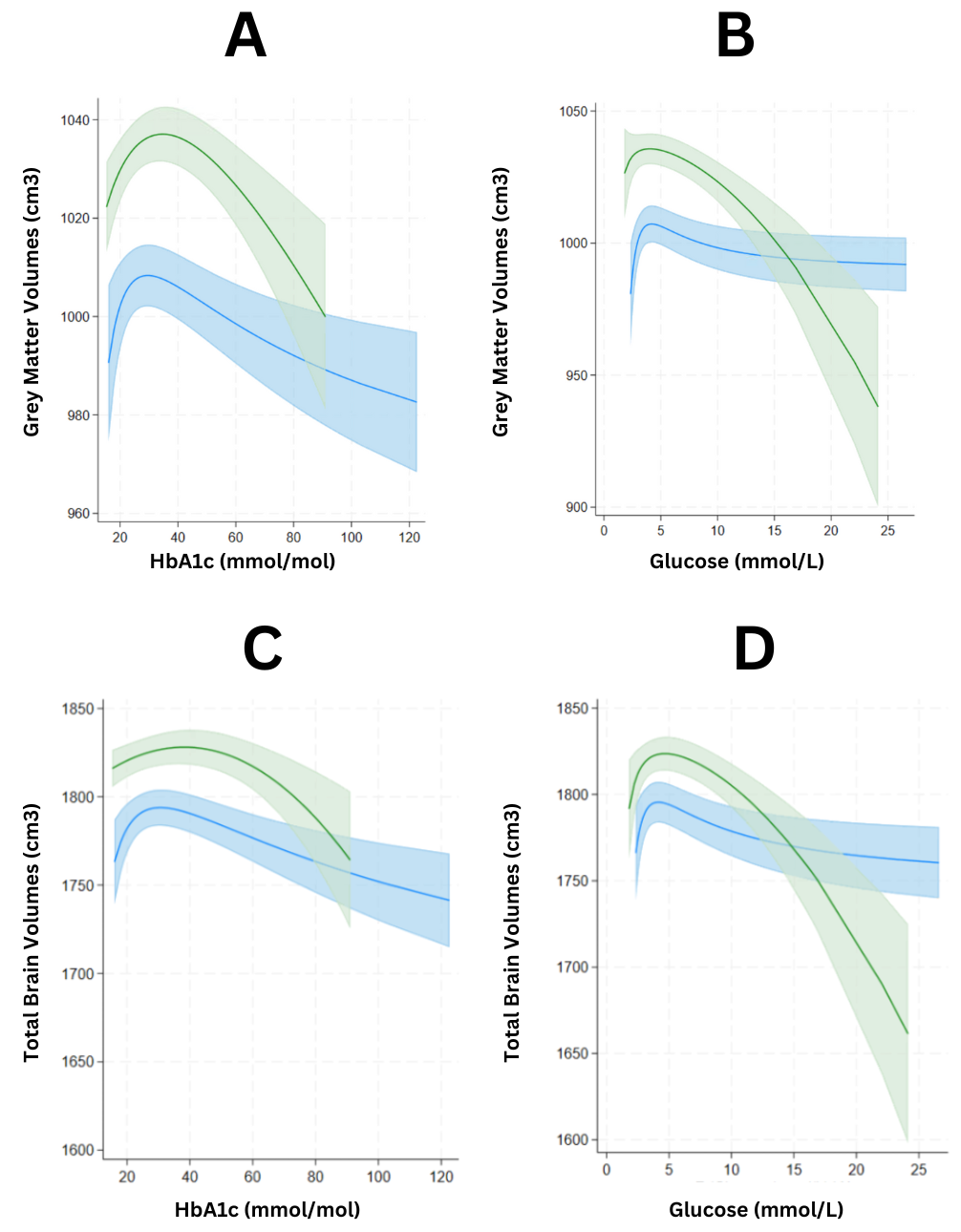

**Supplementary Figure 2:** The partial regression plot of the fractional polynomial models that provided the best fit for the relationship between HbA_1c_/glucose and grey matter volume (A and B) and whole brain volume (C and D). The models presented are the fully confounder-adjusted models excluding participants with diabetes

**Linear models of the straight lines:**

**Methods**: Linear regression models were used to estimate sex-specific associations between glycaemic markers and brain structure within inflection ranges identified from the fractional polynomial models. Models were adjusted for the same sociodemographic and clinical covariates as the main fractional polynomial analyses (Model 2).

**Results:** In linear models restricted to participants with high glucose levels (>10 mmol/L), there was evidence of sex interaction for both whole-brain and grey-matter volume (p_interaction=0·05 and 0·04, respectively), suggesting steeper declines among women compared with men. In women, higher glucose was associated with lower whole-brain (b=−7·22 [95% CI −14·1, −0·34], p=0·04) and grey-matter volumes (b=−4·1 [−8·2, −0·1], p=0·04), whereas associations in men were weaker and imprecise. No evidence of sex interaction was observed for WMHV (p_interaction=0·9).

At comparable HbA1c levels (> 60 mmol/mol), sex interactions were not significant across any brain measure (WBV p = 0·5; GM p = 0·3; WMHV p = 0·1). Nevertheless, women showed a modest positive association between HbA1c and white-matter-hyperintensity volume (b = 0·03 [95% CI 0·001, 0·05], p = 0·03), whereas no effects were observed in men.

|  |  |  | **WBV** | | | **GM** | | | **WMHV** | | |
| --- | --- | --- | --- | --- | --- | --- | --- | --- | --- | --- | --- |
|  |  |  | b | 95% | p | b | 95% | p | b | 95% | p |
| **Glucose** | Males (n = 104) | >10 mmol/L | -1·74 | (-6·4, 2·9) | 0·46 | 0·02 | (-2·5, 2·6) | 0·91 | -0·007 | (-0·07, 0·05) | 0·82 |
|  | Females (n = 42) | >10 mmol/L | -7·22 | (-14·1, -0·34) | 0·04 | -4·1 | (-8·2, -0·1) | 0·04 | -0·01 | (-0·09, 0·07) | 0·74 |
| **HbA1c** | Females (n = 72) | >60 mmol/mol | -0·4 | (-2·2, 1·36) | 0·61 | -0·6 | (-1·2, 0·5) | 0·32 | 0·03 | (0·001, 0·05) | 0·03 |
|  | Males (n = 134) | >60 mmol/mol | 0·2 | (-0·7, 1·2) | 0·68 | 0·1 | (-0·5, 0·7) | 0·73 | 0·1 | (-0·48, 0·71) | 0·71 |

**Supplementary Table 2: Sex-specific linear associations between high glucose or HbA1c levels and brain structural volumes adjusted for the same confounders as the main analyses** (Linear models restricted to high ranges: glucose > 10 mmol/L, HbA1c > 60 mmol/mol)

**Across the lower (“uptick”) range of the exposure–response curve, linear associations were generally small and imprecise for glucose, with little evidence of sex interaction. For HbA1c in the uptick range (20–42 mmol/mol), there was evidence of sex interaction for both hippocampal volume (p_interaction<0·001) and WMHV (p_interaction=0·018).**

**Restricted cubic splines:**

**Methods**: We modelled the association between glycaemic markers and brain volume measures using restricted cubic splines with five knots (placed at the 5th, 27.5th, 50th, 72.5th, and 95th percentiles). Models were adjusted for the same sociodemographic and clinical covariates as the main fractional polynomial analyses (model 2) and included sex×exposure spline interaction terms.

**Results**: Restricted cubic splines supported non-linearity for glucose with WBV (p_non-linearity=0·006) and GM (p_non-linearity<0·001), with evidence of sex interaction for both outcomes (p_interaction=0·02 and 0·03, respectively). For HbA1c, spline models supported non-linearity for WBV (p_non-linearity=0·006) and GM/WM (both p_non-linearity<0·001), with little evidence of sex interaction (p_interaction≥0·4). For WMHV, spline models supported non-linearity for HbA1c (p_non-linearity=0·002) but not glucose, with no evidence of sex interaction for either marker. For HV, spline models showed no evidence of non-linearity for either marker; however, there was evidence of sex interaction for HbA1c–HV (p_interaction=0·0001) but not glucose–HV (p_interaction=0·79).

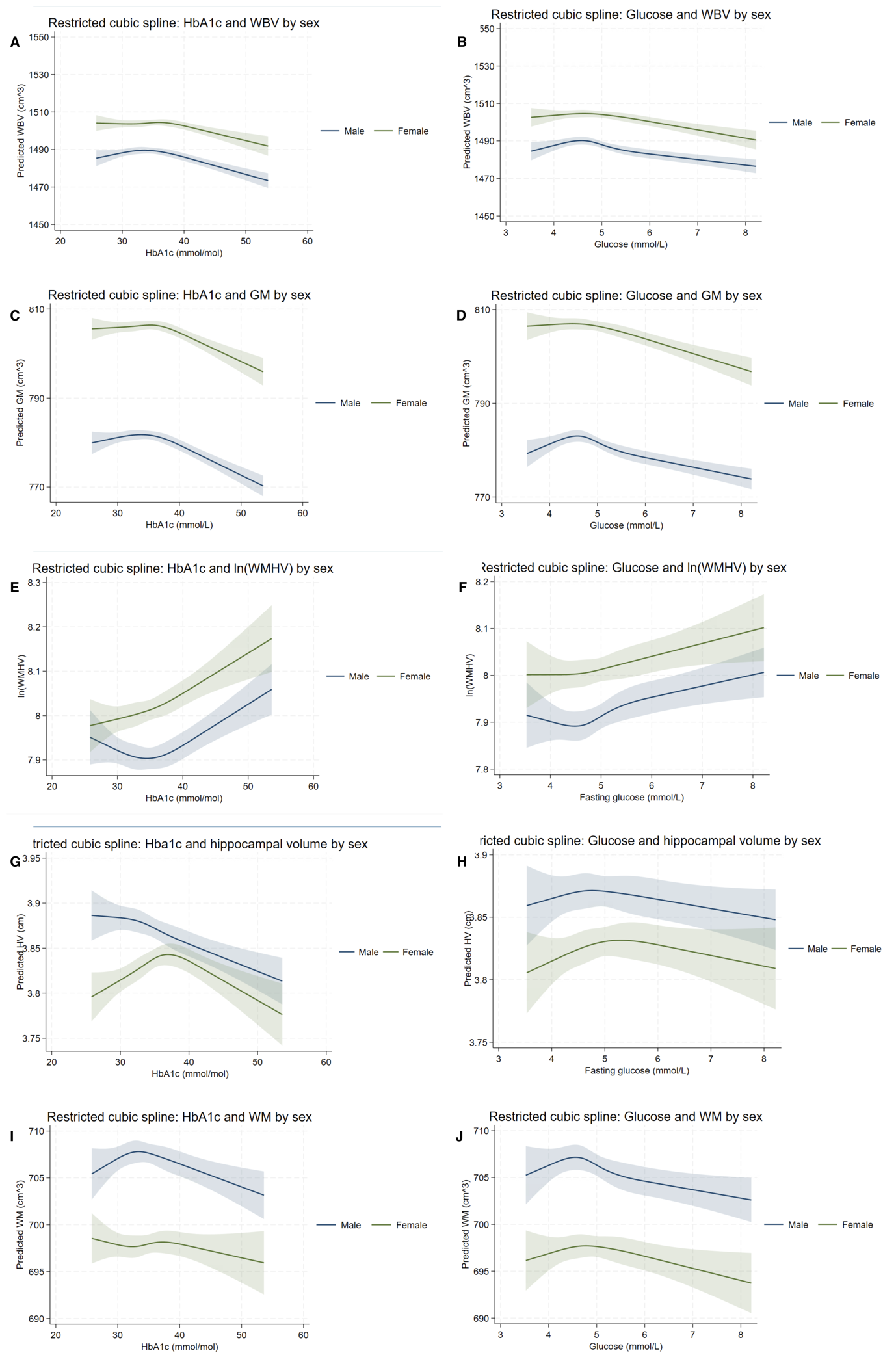

**Supplementary Figure 3. Restricted cubic spline associations of glycaemia with brain MRI volumes.**
Panels show restricted cubic spline (RCS) models of HbA1c and random glucose with MRI-derived brain measures: (A) HbA1c–whole brain volume (WBV), (B) glucose–WBV, (C) HbA1c–grey matter volume (GM), (D) glucose–GM, (E) HbA1c–white matter hyperintensity volume (WMHV; log-transformed), (F) glucose–WMHV (log-transformed), (G) HbA1c–hippocampal volume (HV), (H) glucose–HV, (I) HbA1c–white matter volume (WM), and (J) glucose–WM.

**Peak derivatives:**

To further characterise curve shape and quantify non-linear sensitivity (for the relationships we deemed to be based on the fractional polynomial models [namely WBV, GM and WMHV], we extracted spline-derived **peak derivative coordinates.** For each model (by sex and brain measure), we identified the point of maximum slope—the glycaemia value at which the first derivative of the spline function was greatest in magnitude. This represents the range where predicted brain volume (or WMHV) was most responsive to glycaemic change. Bootstrap resampling (1,000 iterations) was used to obtain bias-corrected standard errors and 95% confidence intervals for each coordinate. The results are reported in the main manuscript.
